# Genetic neurodevelopmental clustering and dyslexia

**DOI:** 10.1101/2023.10.04.23296530

**Authors:** Austeja Ciulkinyte, Hayley S Mountford, Pierre Fontanillas, 23andMe Research Team, Timothy C Bates, Nicholas G Martin, Simon E Fisher, Michelle Luciano

## Abstract

Dyslexia is a learning difficulty with neurodevelopmental origins, manifesting as reduced accuracy and speed in reading and spelling despite adequate education. Dyslexia is substantially heritable and frequently co-occurs with other neurodevelopmental conditions, particularly attention deficit-hyperactivity disorder (ADHD). The purpose of this paper was to elucidate how genetic factors predisposing to dyslexia correlate with risk for other neurodevelopmental and psychiatric traits. A large-scale genome-wide association study (GWAS) of dyslexia diagnosis self-report (51,800 cases and ∼1.1 million controls), together with GWAS of ADHD, autism, Tourette syndrome, anxiety, depression, schizophrenia, bipolar, obsessive compulsive disorder, anorexia, were analysed using Genomic Structural Equation Modelling (GenomicSEM) to construct a genomic structural model. The final model consisted of five correlated latent genomic factors described as F1) internalising disorders, F2) psychotic disorders, F3) compulsive disorders, F4) neurodevelopmental conditions, and F5) attention and learning difficulties, which includes ADHD and dyslexia. This latent factor was moderately correlated with internalising disorders (.40) and, to a lesser extent, with neurodevelopmental conditions (.25) and psychotic disorders (.17), and negatively with compulsive disorders (-.16). Unlike ADHD, most of the genomic variance in dyslexia was unique, suggesting a more peripheral relation to psychiatric traits. We further investigated genetic variants underlying both dyslexia and ADHD. This implicated 49 loci (40 of which were not reported in GWAS of the individual traits) mapping to 174 genes (121 not found in GWAS of individual traits). Our study has discovered novel pleiotropic variants and confirms via GenomicSEM the heightened genetic relation between dyslexia and ADHD versus other psychiatric traits. In future, analyses including additional co-occurring traits such as dyscalculia and dyspraxia, for which there are currently no large-scale GWAS, will allow a more clear definition of the attention and learning difficulties genomic factor, yielding further insights into factor structure and pleiotropic effects.

## Introduction

Dyslexia is classed as a specific learning disorder in the DSM-V ^1^ and is defined by persistent difficulty with accurate and/or fluent word reading and poor spelling ability ^2^. While there are no universal diagnostic criteria, dyslexia is typically identified when reading and writing abilities fall below expectations, considering the individual’s age, exposure to effective education and other cognitive abilities ^3^. Dyslexia is typically identified in childhood, but persists throughout adulthood ^3^. It has been viewed as a neurodevelopmental disorder, linked to structural, connective and functional abnormalities in brain regions involved in visual and auditory processing ^4^. Here, we refer to it as a specific learning difficulty in line with the 2009 Rose Report on dyslexia and reading difficulties ^2^.

Dyslexia is present in 5-10% of children worldwide ^5, 6^, and is the most common specific learning difficulty. It frequently co-occurs with other neurodevelopmental differences—in particular, 25-40% of individuals with dyslexia are diagnosed with attention deficit-hyperactivity disorder (ADHD) and vice versa ^7, 8^. Associations between autism (AUT) and dyslexia are complex. Certain traits, such as atypical sensory processing and spatial attention alterations, are shared between autism and dyslexia ^9^. Yet, some studies show that autism is linked to better reading skills ^10–12^, and others suggest dyslexia is no more prevalent among autistic individuals than in the general population ^13^.

Twin studies of dyslexia estimate its heritability at 60-70% ^14, 15^, which suggests a substantial genetic component. However, the genetic background of dyslexia is complex and multifactorial: individual genes contributing to dyslexia have only a small effect each, and likely act together in an additive manner ^16^. Discovery of such genes requires very large sample sizes, thus previous genome-wide association studies (GWAS) have struggled to identify genomic loci predisposing to dyslexia due to low statistical power ^17^. Through collaboration with the personal genetics company 23andMe, Inc, Doust and colleagues published the largest dyslexia GWAS to date, comprising over 1.1 million individuals in total (∼50,000 dyslexia cases) and discovering 42 significantly associated genomic loci ^18^. This dataset enables further study of the genetic background of dyslexia.

Genetic factors underlying neurodevelopmental and psychiatric traits often overlap between disorders: of 208 genes associated with at least one psychiatric disorder, it was reported that approximately half of them are also associated with another disorder ^19^. Recent developments in structural equation modelling methods to study multiple phenotypes with overlapping genomic influences permit quantitative analysis of genetic correlations between individual psychiatric traits/disorders ^20^. Such studies aim to construct a structural model where traits are clustered based on their genetic similarity (using the correlational structure between genome-wide single-nucleotide polymorphism (SNP) associations for each trait that give rise to genetic correlations). Each cluster is described by a single latent factor, which represents the shared genetic (SNP) liability within the cluster. In 2019, structural modelling of 8 psychiatric disorders proposed a three-factor model describing three clusters of genetically correlated disorders ^21^, while a 2022 follow-up analysis of 11 psychiatric disorders proposed a four-cluster model ^22^. Broadly, these clusters are: 1) Early-onset neurodevelopmental disorders: ADHD, autism, Tourette syndrome, major depressive disorder; 2) Disorders with compulsive behaviours: obsessive-compulsive disorder, anorexia nervosa, Tourette syndrome; 3) Mood and psychotic disorders: bipolar disorder, schizophrenia, (major depressive disorder in three-factor model), and; 4) Internalising disorders (four-factor model): anxiety disorder, major depressive disorder.

To understand whether and where dyslexia is located among these broad genetic clusters, here we expand and adapt current genomic structural models to include dyslexia. Based on prior literature, we expected that dyslexia would fall under an early-onset neurodevelopmental disorder factor. However, we made no prediction of how correlated dyslexia would be with this neurodevelopmental factor given that the nature of the latent factor itself depends on the range of variables included in the analysis. For instance, an earlier GenomicSEM analysis of 8 traits, identified a neurodevelopmental factor as loading on Tourette Syndrome, major depressive disorder, ADHD and autism ^21^. However, a later study ^22^ adding problematic alcohol use, post-traumatic stress disorder (PTSD), and anxiety found that the neurodevelopmental factor also loaded on problematic alcohol use and PTSD, while the loading onto major depressive disorder on this factor reduced from .60 (in the model with 8 traits) to just .20. In another study ^23^, addition of alcohol dependence, nicotine dependence and cannabis use disorder to the original 8 traits resulted in a neurodevelopmental factor that loaded more strongly on major depression than any of the developmental disorders, and that had strong loadings on alcohol and nicotine dependence, thus changing the nature of the factor.

In the present study, we focussed on 10 developmental/psychiatric traits and expected to observe a four latent factor model—compulsive, psychotic, neurodevelopmental, and internalising—with dyslexia clustering especially with ADHD for which it is known to be moderately genetically correlated (*r* = .53) ^18^. Because our aim was to more clearly delineate neurodevelopmental genomic factors influencing phenotypes expressed in childhood irrespective of environment we excluded substance use dependence and PTSD due to these trait’s dependence on environmental exposures Additionally, because neurodevelopmental traits are linked to social outcomes, they may themselves create environments that then influence the expression of other traits that do not have origins in neurodevelopment (e.g., a child with dyslexia may disengage with school, develop low self-esteem or feel alienated ^24^, putting them at greater risk of substance use ^25^) but through a causal pathway would correlate with a neurodevelopmental factor. We followed up our analysis with targeted investigations of genomic loci associated with both dyslexia and ADHD, to provide the strongest evidence thus far of pleiotropic effects.

## Methods

### Samples

To construct the genomic structural model, we sourced publicly available GWAS summary statistics for 10 neurodevelopmental/psychiatric traits (Table 1). GWAS summary statistics for all traits, except dyslexia, were obtained from multi-cohort case-control meta-analyses. The dyslexia summary statistics came from a single analysis of 23andMe, Inc, participants in which genomic inflation was controlled. The combined sample amounted to 453,408 cases and 2,374,026 controls, and included some sample overlap, for example, iPSYCH was part of the ADHD, ASD, and BPD GWAS. Consistent with previous genomicSEM investigations, data were restricted to participants with European ancestry as these currently have adequate sample sizes.

**Table 1.** Sources and description of GWAS summary statistics used for GenomicSEM modelling.

| **Disorder** | **Abbreviation** | **N(cohorts)** | **N(cases)** | **N(controls)** |
| --- | --- | --- | --- | --- |
| Attention deficit/hyperactivity disorder <sup>32</sup> | ADHD | 13 | 38,691 | 186,843 |
| Anorexia nervosa <sup>67</sup> | AN | 33 | 16,992 | 55,525 |
| Anxiety disorder <sup>68</sup> | ANX | 2 | 53,978 | 221,844 |
| Autism spectrum disorder <sup>69</sup> | AUT | 6 | 18,381 | 27,969 |
| Bipolar disorder <sup>70</sup> | BIP | 57 | 41,917 | 371,549 |
| Dyslexia <sup>18</sup> | DYX | 1 | 51,800 | 1,087,070 |
| Major depressive disorder <sup>71</sup> | MDD | 3 | 170,756 | 329,443 |
| Obsessive-compulsive disorder <sup>72</sup> | OCD | 2 | 2,688 | 7,037 |
| Schizophrenia <sup>73</sup> | SCZ | 90 | 53,386 | 77,258 |
| Tourette syndrome <sup>74</sup> | TS | 4 | 4,819 | 9,488 |

### Data standardisation and quality control

To ensure all data were uniform and reliable, all GWAS summary data were aligned to the 1000 Genomes European reference genome build 37 ^26^ and filtered to imputation quality score >0.9, minor allele frequency >0.05 using the *sumstats* and *munge* functions in the GenomicSEM R package ^27^. Any SNPs not commonly shared between all 10 studies were excluded. After quality control, 3,959,995 SNPs remained for further analysis.

### Statistical analysis

#### GenomicSEM

SNP-based heritability and pairwise genetic correlations (r_g_) between disorders were obtained using the linkage disequilibrium score regression (LDSC) ^28^ function in the GenomicSEM package and based on 830,359 high quality HapMap SNPs. To reveal clusters of traits with shared genetic liability, we synthesised LDSC outputs into a genomic structural model. Our initial model was guided by a previously published three latent factor structural model of 8 psychiatric disorders ^21^, all of which were also included in the present study. Given that anxiety and dyslexia were new to this model, we further investigated the factor structure underlying these genetic relationships using an exploratory factor analysis (EFA) of the genetic covariance matrix with *promax* rotation. Goodness of fit of the confirmatory and exploratory models was evaluated by the standard fit statistics using recommended criteria: lower Akaike’s Information Criterion (AIC), Comparative Fit Index (CFI) in the range of .97 and 1 (good; .95-.97, acceptable), and Standardised Root Mean Squared Residual (SRMR) < .05 (good; .05-.1, acceptable) ^29^.

#### Identification of common dyslexia and ADHD variants

Given the frequent co-occurrence of ADHD and dyslexia and their strong genetic correlation, we sought to discover pleiotropic genetic loci that are significantly associated with both traits. We filtered dyslexia and ADHD datasets to 3,956,700 shared SNPs and calculated an overall effect size (r) and degree of sharedness (Θ) for each SNP using a polar coordinate transformation method PolarMorphism ^30^. By taking Θ into account, we were able to correct for inflation in effect size due to correlation and sample overlap ^30^. SNPs where FDR-adjusted p-values (q-value) for r and Θ were <0.05 were deemed significant and brought forward for gene mapping. Functional annotation and gene mapping was performed using FUMA ^31^. We clumped linkage-independent genomic regions (r^2^ threshold = 0.4, maximum LD distance = 500 kb, maximum p-value for lead SNPs = 5 × 10^-8^, maximum p-value cut-off = 5 × 10^-2^). The MHC region was considered as one locus. The 1000 Genomes European population was used as reference (GRCh37 release).

## Results

### Genetic Clustering

Heritability Z-scores were >4, LDSC intercepts approximately 1, and the ratio close to 0 indicating that linkage disequilibrium scores reflected polygenic heritability. The strongest genetic correlations were observed between anxiety (ANX) and major depressive disorder (MDD) (r_g_ = 0.86 ± 0.05), then bipolar disorder (BIP) and schizophrenia (SCZ) (r_g_ = 0.69 ± 0.03), with moderate correlations ranging between 0.40 and 0.45 for pairings of ANX with ADHD and BIP, for MDD with ADHD, BIP and SCZ, for anorexia nervosa (AN) and obsessive-compulsive disorder (OCD), and ADHD and dyslexia (DYX) (see Figure 1A and B). A regression of effective sample size on estimated genetic correlation for each pair of disorders indicated that there were no effects of sample size on genomic correlation (R^2^_adj_ = –0.02, p = 0.90, Figure 1C).

**Figure 1.**
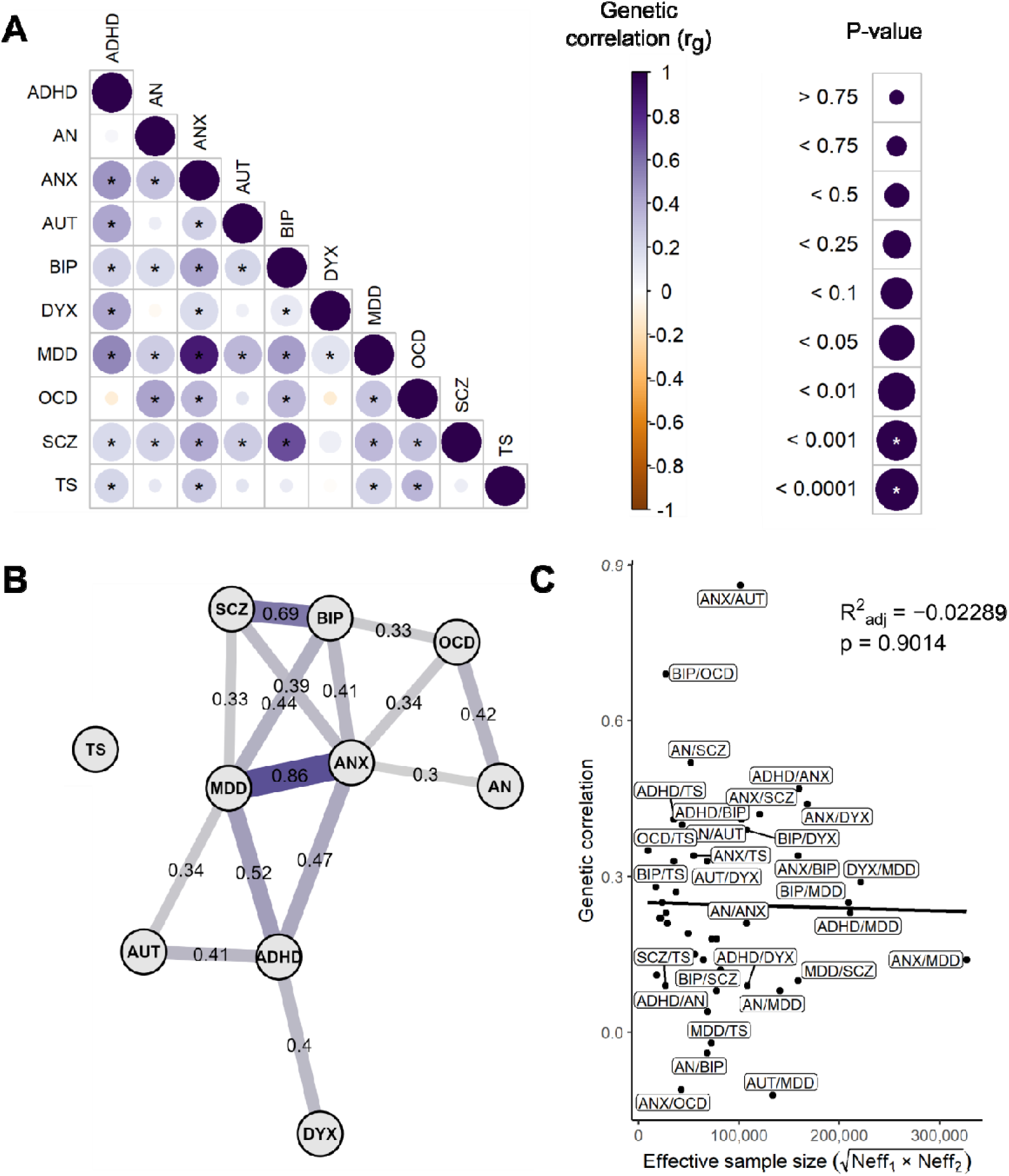
Genetic relationships between ten neurodevelopmental and psychiatric disorders. **A** – pairwise genetic correlations detected using LDSC. Colour intensity scales with correlation coefficient (r_g_), radius of circles scales with significance of p-values. Asterisks denote statistically significant (p ≤ 0.001) correlations after Bonferroni correction. **B** – path diagram of genetic correlations. Each edge connecting two phenotype nodes represents genetic correlation between those traits. Width and colour intensity of edges scale with correlation coefficient (r_g_). Only pairs where r_g_ > 0.3 and correlation is statistically significant (p ≤ 0.05) after Bonferroni correction are displayed. **C** – regression of effective sample size on estimated genetic correlation for each pair of traits. Pairs where r_g_ > 0.3, or r_g_ < 0.1, or effective sample size >100,000 are labelled.

Due to their strong correlation, we modelled ANX to load on the same two latent factors as MDD. Similarly, we modelled DYX to load on the same factor as ADHD. However, the fit of this model was poor (χ^2^(29) = 329.40, AIC = 381.40, CFI = 0.933, SRMR = 0.077). An EFA identified a structure of four correlated factors, which together explain 61.3% of underlying variance. The model identified by EFA clusters ANX and MDD under a common factor of internalising disorders. This is in line with the newer report of an 11-disorder structural model^22^. We thus constructed a confirmatory model using the EFA output (Figure 2A) and observed a dramatic improvement in model fit (χ^2^(29) = 172.60, AIC = 224.60, CFI = 0.968, SRMR = 0.062). Having observed a strong genetic correlation between DYX and ADHD but not between DYX and AUT, we modified the confirmatory model by modelling DYX and ADHD to load on a fifth factor (F5) of learning difficulties (Figure 2B). This model had the best fit of all the estimated models χ^2^(24) = 96.57, AIC = 158.57, CFI = 0.984, SRMR = 0.048).

**Figure 2.**
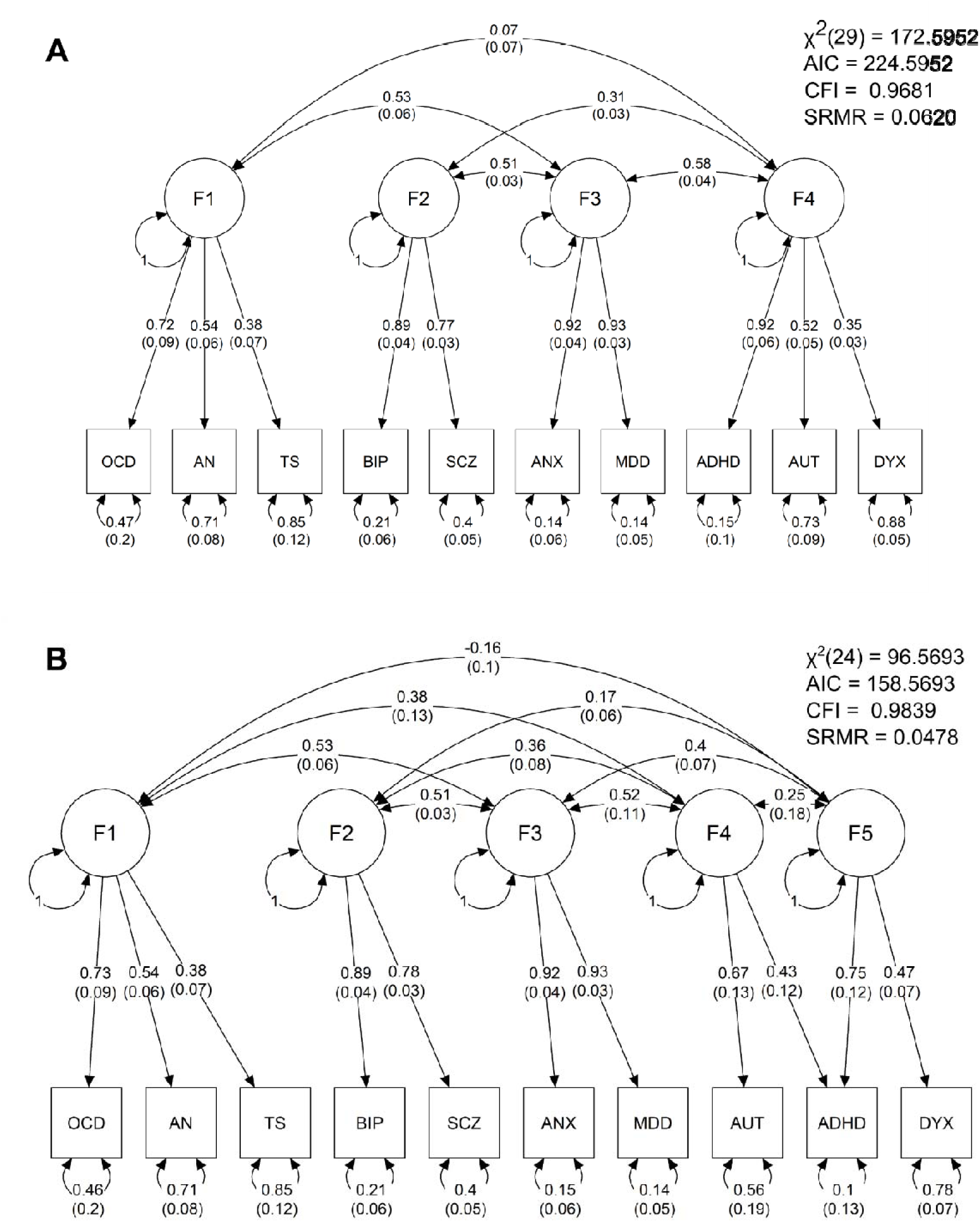
Structural models of 10 neurodevelopmental and psychiatric disorders. Each genetic factor (F1 – F5) represents shared genetic liability. Single-headed arrows represent standardised loading parameters, which indicate covariance of the latent factor with a given parameter. Standard errors are given in parentheses. Double-headed arrows connecting factors represent pairwise correlation. Double-headed arrows connecting a component to itself represent residual variance, i.e., variability that is unexplained by factor loading. Factor residuals are fixed for scaling. **A** – confirmatory model based on exploratory factor analysis. **B** – modified confirmatory model that separates DYX and ADHD into a separate cluster of learning difficulties, based on observed genetic correlations.

### Identification of shared variants contributing to dyslexia and ADHD

From the overlapping SNPs available for DYX and ADHD, 1,566 SNPs met the significance criteria for overall effect size and degree of sharedness. These 1,566 pre-defined significant SNPs were clumped into 51 lead SNPs belonging to 49 genomic risk loci (Figure 3A). Overall and standardised genomic inflation factors were close to 1 (λ = 0.879, λ_1000_ = 0.999), indicating that bulk inflation and excess false positive rates were minimal (Figure 3B). MAGMA tissue expression analysis indicated that these SNPs are associated with genes showing enriched expression in brain tissues (Figure 3C). Six out of 49 pleiotropic loci were previously reported as associated with dyslexia ^18^, and one of these 6 (lead SNP rs1005678 on chromosome 3) was also found in the ADHD GWAS ^32^. A further 3 of the pleiotropic loci were reported for ADHD alone ^32^ (Figure 3D). Forty-nine pleiotropic loci were mapped to 174 protein coding genes (Supplementary Table 3). Gene Ontology analysis ^33–35^ indicated an enrichment in genes involved in protein modification and metabolism, and in development (Table 2). Thirty-six out of the 174 pleiotropic protein-coding genes have been previously associated with dyslexia (i.e., from 173 genes mapped to the 42 significant loci from the dyslexia GWAS) ^18^, and 21 with ADHD ^32^. Of those, four genes, *TCTA* (T-cell leukemia translocation altered), *AMT* (aminomethyltransferase), *TRAIP* (TRAF interacting protein) and *SORCS3* (sortilin-related receptor 3) had been associated with both traits in prior literature (Figure 3E).

**Figure 3.**
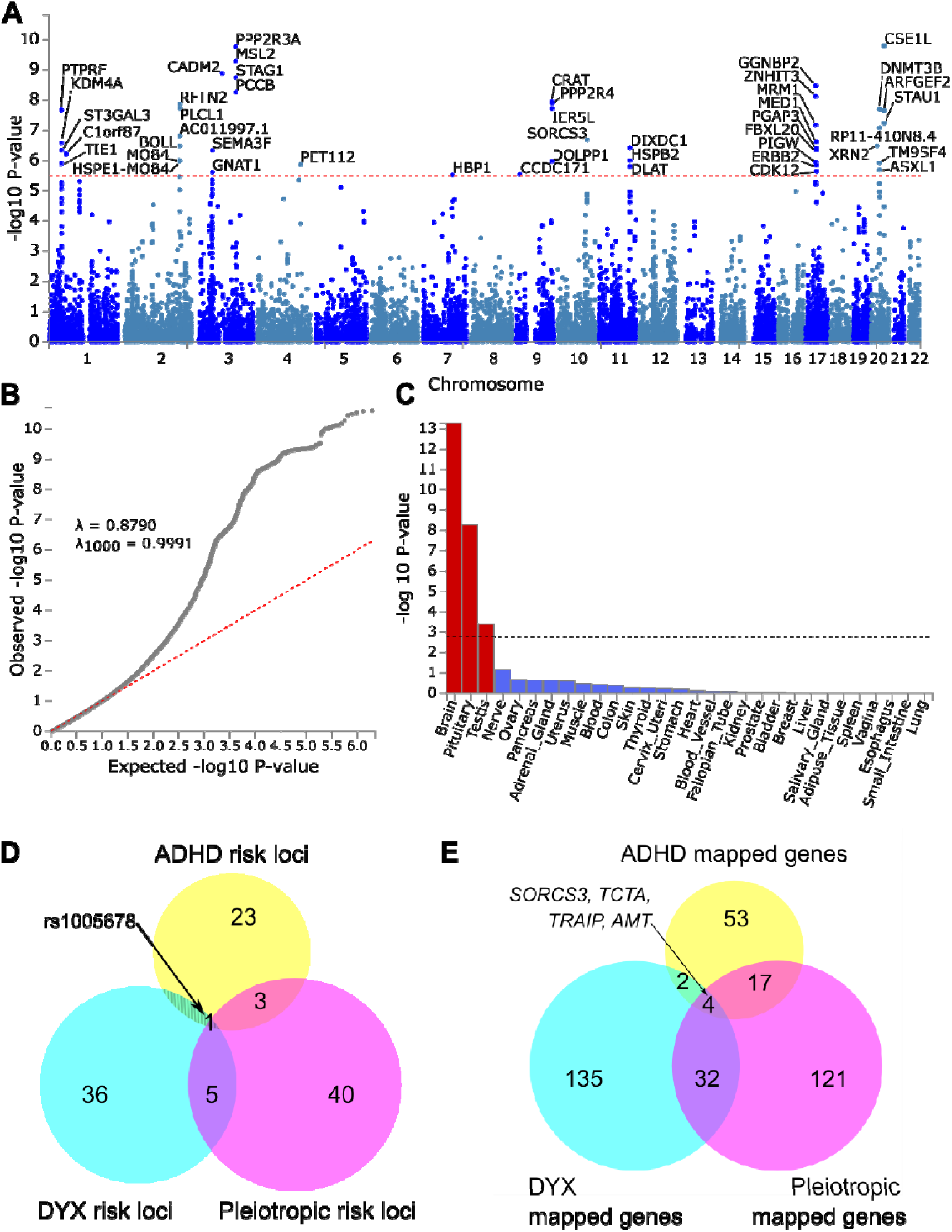
Investigating pleiotropic genomic loci influencing dyslexia and ADHD. **A** – Manhattan plot of pleiotropy effect size p-values across SNPs shared between dyslexia and ADHD GWAS datasets. Dotted line represents Bonferroni-significant p-value (3.081 × 10^−6^). **B** – Quantile-quantile plot displaying the observed vs expected statistics under the null hypothesis. **C** – MAGMA tissue expression analysis of all SNPs using GTEx v8 dataset with 30 general tissue types. Dotted line represents Bonferroni-significant p-value. **D** – Venn diagram of significantly associated genomic loci identified in single phenotype GWAS studies and in the pleiotropy analysis. **E** – Venn diagram of genes mapped to significantly associated SNPs in single phenotype GWAS studies and in the pleiotropy analysis.

**Table 2.**
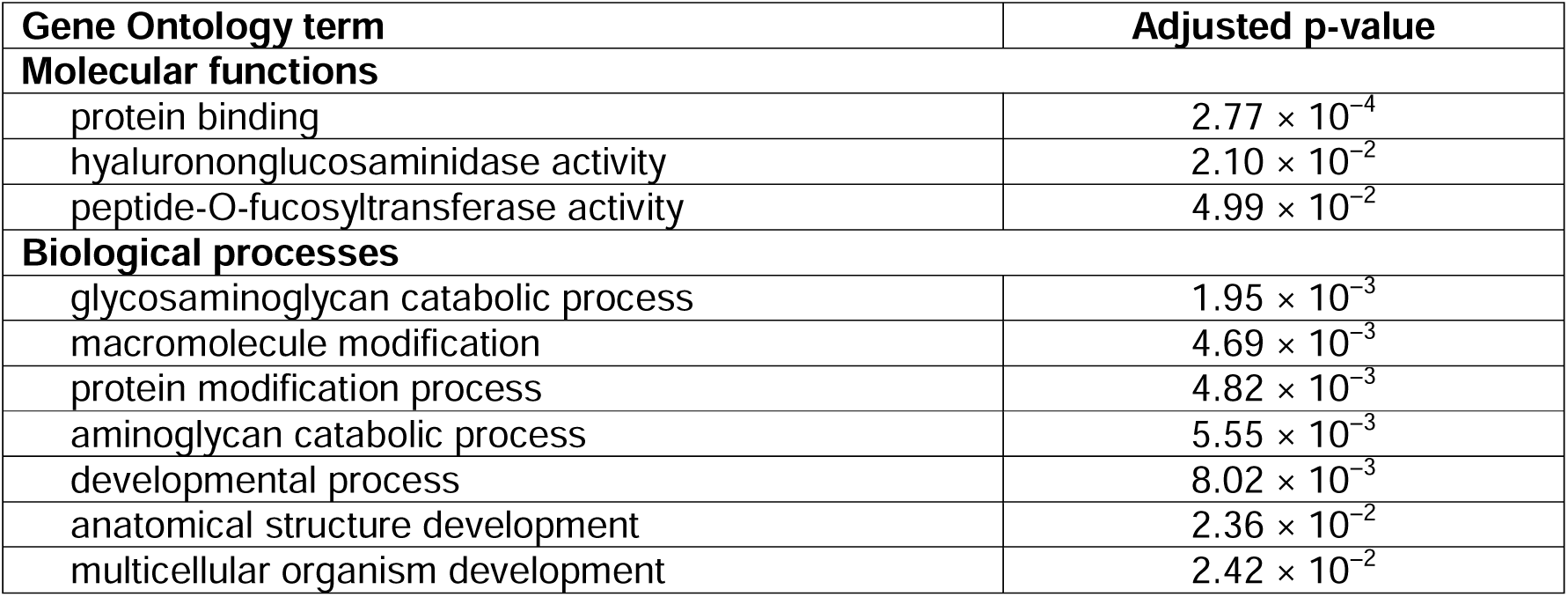
Results of Gene Ontology enrichment analysis for 174 genes mapped to putative pleiotropic loci influencing dyslexia and ADHD.

## Discussion

By extending existing multivariate genomic models of neurodevelopmental and psychiatric traits to include dyslexia, our work has yielded three key findings: (1) that a five latent factor model, composed of internalising (F1), psychotic (F2), compulsive (F3), neurodevelopmental (F4), and attention and learning latent traits (F5), effectively describes the genetic relationships between these diagnoses; (2) that ADHD aligns more with dyslexia and to a learning difficulties latent factor than a neurodevelopmental one, and; (3) we identify a set of pleiotropic genetic loci associated with the presence of both dyslexia and ADHD.

Genetic correlations between the psychiatric disorders were concordant with those of previous analyses ^21, 22^. Most pairs of disorders displayed a statistically significant genetic correlation, varying in magnitude, as observed previously. This supports the concept of a complex, interlinked network of shared genetic liabilities across psychiatric disorders. The moderate genetic correlation (0.40) between dyslexia and ADHD matches estimates derived from a meta-analysis of twin studies for reading ability indicators and ADHD symptoms ^36^. This result was expected, given the frequent co-occurrence of ADHD and dyslexia ^7, 37^, correlation between general reading ability and ADHD ^36, 37^, and significant ADHD polygenic score prediction of dyslexia and reading achievement ^17, 32^.

Based on the frequent co-occurrence of dyslexia and ADHD, and, albeit less often, with autism, we initially hypothesised a structural model that includes dyslexia alongside other neurodevelopmental disorders. However, genetic correlation between dyslexia and autism was found to be statistically non-significant (p = .082), and accordingly, an improvement in model fit was observed when dyslexia and autism were modelled as loading on distinct latent factors. By contrast with dyslexia, ADHD loaded on both the neurodevelopmental and the attention and learning difficulties factors (F4 and F5), with its correlation being lower on the former than on the latter (.43 vs .75). This suggests that the genetic architecture influencing ADHD shows greater overlap with dyslexia than with autism. We propose four non-mutually exclusive explanations: (a) ADHD is a complex, biologically heterogeneous disorder that shares genetic influences with both neurodevelopmental and learning difficulties to varying degrees; (b) vertical (or spurious) pleiotropy is present and may reflect causal influences of ADHD on dyslexia or vice-versa; (c) current diagnostic criteria for dyslexia, ADHD and autism are insufficient to categorically define genetically distinct populations, and suggest different degrees of overlap between the continuous traits underlying the diagnoses, and; (d) co-occurrence of dyslexia is higher than autism within the GWAS sample ascertained for ADHD ^32^. Although the PolarMorphism method used corrects for vertical pleiotropy and potential sample overlap ^30^.

The shared and unique genetic architecture identified in this study is broadly in line with previous genomic structural models ^20–22^ and highlights the complexity of behavioural and psychiatric genetics. We observed strong positive correlations between the five latent factors in our structural model, which supports the concept of general genetic risk factors that contribute to all 10 neurodevelopmental and psychiatric traits in this panel. However, we also observe a range of residual variances, which represent specific genetic factors that confer distinguishable phenotypes to each developmental trait/psychiatric disorder. Dyslexia and autism have larger residual estimates than ADHD (0.78, 0.56 and 0.10, respectively). This suggests that (a) shared genetic factors explain a larger part of total genetic influence for ADHD than for either dyslexia or autism; (b) both dyslexia and autism have a substantial proportion of risk factors not shared with ADHD that pertain more specifically to biological mechanisms related to these traits’ core features (i.e., respective reading subskills and social communication). Additionally, a recent bivariate causal mixture model analysis of bipolar disorder, depression, schizophrenia, and ADHD showed that ADHD is the least polygenic of these traits (5600 causal variants) ^38^. If polygenicity is low but pleiotropy high then one would expect lower residual variance for ADHD. Thus, the finding of substantial residual variance in dyslexia and autism could indicate higher polygenicity for these traits compared to ADHD. Intelligence, a trait genetically correlated with dyslexia, ADHD, and autism, for instance, showed high polygenicity (∼11,500 variants) using this mixed effect modelling ^38^. Such an approach should be applied to dyslexia and autism in future.

We observed that the neurodevelopmental latent factor was more strongly correlated than the attention and learning factor with the other latent factors (often showing correlations that were twice as high). This aligns with findings from a study that dissected the shared and unique genetic background of ADHD and autism spectrum disorder ^39^. That study found that the shared genomic portion was strongly associated with psychiatric traits (e.g., depressive symptoms) whereas the distinct part was strongly associated with cognitive traits (e.g., educational attainment, childhood IQ). This distinct part is likely captured in the attention and learning difficulties factor. Further extension of the genomic structural model including other relevant traits should help to clarify the nature of the attention and learning factor and the large residual variance in dyslexia. Specifically, additional specific learning difficulties, such as dyscalculia, and other developmental disorders that co-occur with dyslexia, namely developmental language disorder and dyspraxia ^40^ should be prioritised for inclusion. At present, no large GWAS have been reported for these traits, precluding their use for GenomicSEM. Multivariate genetic modelling in 12-years old twins ^41^ showed that genetic influences on reading, mathematics, and language difficulties each overlapped largely with the genetic influences on general cognitive ability/low ability, so an analysis that further incorporates general cognitive ability may show that it correlates strongly with the attention and learning difficulties factor.

Our targeted approach to identify pleiotropic loci associated with both dyslexia and ADHD uncovered 49 shared genomic loci. Importantly, 43 of these loci were not among genome-wide significant associations of the prior source GWAS of each separate trait, and thus represent newly identified pleiotropic loci. The 49 putative pleiotropic loci were mapped to 174 protein-coding genes, with gene ontology analysis suggesting enrichment of genes involved in development. This is consistent with the neurodevelopmental origins of both dyslexia and ADHD, both manifesting from changes in structure, connectivity and function of the brain ^42, 43^. We also detected enrichment of genes involved in protein modification and metabolism. While it is known that post-translational modification has a major role in neurodevelopment in general ^44^, any specific links to dyslexia and ADHD require further investigation.

Of the 49 significant SNPs, 13 showed no associations in the GWAS Catalog with primary phenotypes (developmental, cognitive, attainment) related to dyslexia or ADHD. One might place lower confidence in these findings given that variants shared between dyslexia and ADHD would likely have generalised effects that can be detected in related traits such as educational attainment or cognitive function which have well-powered GWAS. Six SNPs were associated with dyslexia in the prior GWAS, with one of these also significant in the previous ADHD GWAS. GWAS Catalog look-up showed a further two of these SNPs associated with ADHD or a combined phenotype including ADHD, and two others with the related traits of educational attainment and cognitive function (including processing speed). There were no reported associations with relevant traits for rs73175930 in *AUTS2*. However, ADHD is a core feature of individuals with AUTS2 syndrome arising from pathogenic variants in this gene ^45^. For the three SNPs previously identified in the ADHD GWAS but not the dyslexia GWAS, all were reported to associate with educational attainment and/or cognitive function. For all other significant SNPs, there was a mixture of reported associations with primary traits, secondary traits (risk taking, externalising behaviours, psychiatric, neuroticism), and other traits (many medical health outcomes that may be downstream outcomes related to lower SES of those with dyslexia and ADHD). These variants may therefore be associated with a whole range of behaviours due to their primary effect on attention and learning processes which we suggest define the covariation between dyslexia and ADHD. This hypothesis might be further tested in extended genomicSEM models that include a host of variables that we identify here (e.g., risk-taking, externalising behaviours, educational attainment) as being previously associated with our significant SNPs for combined dyslexia and ADHD. Downstream outcomes can be confirmed by Mendelian Randomization methods in cases where confounding by socio-economic status is unproblematic.

Four genes—*SORCS3, TCTA, TRAIP and AMT*—shown to be pleiotropic had previously been associated with dyslexia and ADHD in individual GWAS studies and are very strong pleiotropic candidate genes ^18, 32^. The *SORCS3* protein is abundant in the central nervous system ^4646^. It has a primary role in sorting intracellular proteins between organelles and the plasma membrane, and a secondary role in cell signalling. Mouse studies of the murine orthologue of *SORCS3* have implicated it in long-term synaptic depression via aberrant glutamate signalling ^47^. *SORCS3*-deficient mice have decreased synaptic plasticity and deficits in spatial learning and memory. This is consistent with the proposed theories of reduced visual and spatial learning abilities in children with dyslexia and ADHD ^48, 49^. *SORCS3* has previously been suggested as a pleiotropic gene associated with ADHD, autism, schizophrenia, bipolar, and MDD ^50^. In addition, *SORCS3* mutations have been linked with intellectual delay ^51^, multiple sclerosis ^52^ and Alzheimer’s disease ^53^. *TCTA* has a role in regulating processes related to dissolution and absorption of bone ^54^, so is not an obvious candidate gene for involvement in brain-related phenotypes. However, in GWAS, variants at this locus have been associated with relevant traits of very high intelligence, cognitive function, and household income ^55–57^, and less relevant traits like cardio-vascular disease, Crohn’s Disease, inflammatory bowel disease (from NHGRI-EBI GWAS Catalog ^58^). *TRAIP,* part of the RING finger protein gene group is linked to the ubiquitination pathway protecting genome integrity following replication stress ^59^. Variants in this gene have been associated with more than 40 phenotypes, but notably the most strongly associated SNPs (P ≤ 7 x 10^-18^) in this gene have been from a meta-analysis combining ADHD, ASD and intelligence ^60^, for multiple studies of intelligence/cognitive function including the cognitive component of education attainment ^61–64^, and externalising behaviour ^65^. Thus, the gene may potentially be involved in general learning processes that can be affected in both ADHD and dyslexia. The *AMT* gene encodes a critical component of the glycine cleavage system contributing to normal development and function of neurons ^66^. It is strongly associated (P = 5 x 10^-82^) with educational attainment ^63^, an outcome that is negatively correlated with both ADHD and dyslexia, and could be prioritised as a candidate gene involved in learning given its lack of strong association with other phenotypes. For the remaining 170 pleiotropic mapped genes (32 previously found for dyslexia and 17 for ADHD), and particularly the 121 that were not identified in previous dyslexia and ADHD screens, future research is needed to understand the extent of their overlap with both general and specific cognitive abilities.

There are a number of limitations to this study: (1) the GWAS included here had variable sample sizes. Lack of power in small GWAS datasets (i.e., AN, TS) results in decreased effect sizes, and thus could contribute to reduced strengths of genetic correlations observed. (2) We have been unable to control for causal relationships and diagnostic overlap in building our structural models, which may potentially inflate genetic correlations.

In sum, our analysis of genetic relationships of 10 developmental traits (which includes dyslexia for the first time) and psychiatric disorders has shown the emergence of an attention and learning difficulties factor that is only modestly correlated with a separate neurodevelopmental factor. In this model, ADHD aligns more closely with dyslexia than autism, suggesting that ADHD may be better termed as a learning difficulty than a psychiatric disorder, and highlighting the importance of it being managed within education and later employment. To explain the large residual variance in dyslexia, extension of the genomicSEM model to include other co-occurring developmental traits and a range of other cognitive abilities will be informative, once reliable GWAS of these are available. Finally, we discovered 49 potentially pleiotropic genomic risk loci, 43 of which are novel, influencing the development of both dyslexia and ADHD, and further confirm *SORCS3* and *TRAIP* as putative pleiotropic genes that likely have broad associations with neuropsychiatric traits potentially through learning pathways. Future GWAS investigations of individuals with co-occurring dyslexia and ADHD will help to validate our pleiotropy analyses and determine whether the identified variant effects are larger when both traits are present.

## Data Availability

The full GWAS summary statistics for the 23andMe discovery data set will be made available through 23andMe to qualified researchers under an agreement with 23andMe that protects the privacy of the 23andMe participants. Datasets will be made available at no cost for academic use. Please visit https://research.23andme.com/collaborate/#dataset-access/ for more information and to apply to access the data.

## Acknowledgements

AC is funded by the Wellcome Trust [218493/Z/19/Z]. HSM is supported by the Biotechnology and Biological Sciences Research Council [BB/T000813/1]. SEF is supported by the Max Planck Society (Germany). The following members of the 23andMe Research Team contributed to this study: Stella Aslibekyan, Adam Auton, Elizabeth Babalola, Robert K. Bell, Jessica Bielenberg, Jonathan Bowes, Katarzyna Bryc, Ninad S. Chaudhary, Daniella Coker, Sayantan Das, Emily DelloRusso, Sarah L. Elson, Nicholas Eriksson, Teresa Filshtein, Pierre Fontanillas, Will Freyman, Zach Fuller, Chris German, Julie M. Granka, Karl Heilbron, Alejandro Hernandez, Barry Hicks, David A. Hinds, Ethan M. Jewett, Yunxuan Jiang, Katelyn Kukar, Alan Kwong, Yanyu Liang, Keng-Han Lin, Bianca A. Llamas, Matthew H. McIntyre, Steven J. Micheletti, Meghan E. Moreno, Priyanka Nandakumar, Dominique T. Nguyen, Jared O’Connell, Aaron A. Petrakovitz, G. David Poznik, Alexandra Reynoso, Shubham Saini, Morgan Schumacher, Leah Selcer, Anjali J. Shastri, Janie F. Shelton, Jingchunzi Shi, Suyash Shringarpure, Qiaojuan Jane Su, Susana A. Tat, Vinh Tran, Joyce Y. Tung, Xin Wang, Wei Wang, Catherine H. Weldon, Peter Wilton, Corinna D. Wong. For the purpose of open access, the author has applied a Creative Commons Attribution (CC BY) licence to any Author Accepted Manuscript version arising from this submission.

## Dyslexia GWAS summary data availability

The full GWAS summary statistics for the 23andMe discovery data set will be made available through 23andMe to qualified researchers under an agreement with 23andMe that protects the privacy of the 23andMe participants. Datasets will be made available at no cost for academic use. Please visit https://research.23andme.com/collaborate/#dataset-access/ for more information and to apply to access the data. Note that 23andMe participants provided informed consent and volunteered to participate in the research online, under a protocol approved by the external AAHRPP-accredited IRB, Ethical & Independent (E&I) Review Services. As of 2022, E&I Review Services is part of Salus IRB (https://www.versiticlinicaltrials.org/salusirb).

